# Epidemiological surveillance of respiratory pathogens in wastewater in Belgium

**DOI:** 10.1101/2022.10.24.22281437

**Authors:** Annabel Rector, Mandy Bloemen, Marijn Thijssen, Bram Pussig, Kurt Beuselinck, Marc Van Ranst, Elke Wollants

## Abstract

**Background:** Detailed information on the circulation of respiratory viruses in the community is crucial to gain better understanding of the burden of respiratory infections on society.

**Methods:** By using an in-house respiratory panel for simultaneous detection of 29 respiratory pathogens (22 viruses and 7 bacteria/fungi), we explored the possibility to use wastewater sampling to monitor the circulation of respiratory pathogens at population level.

**Results:** We were able to detect all respiratory viruses included in the panel (influenza A, respiratory syncytial virus (RSV), human metapneumovirus (HMPV), parainfluenza viruses (PIV) 1-4, adenovirus (Adv), human bocavirus (HBoV), enterovirus/rhinovirus (EV/RV), enterovirus D68 (EV-D68), parechovirus (HPeV), human coronaviruses (HCoV)-NL63, -229E, -OC43, -HKU-1 and -SARS, cytomegalovirus (CMV) and herpes simplex virus (HSV)-1 and -2), except for influenza B and HCoV MERS which were not circulating in Belgium during the two year study period. An upsurge of EV-D68 infections in Europe in September 2021 was clearly reflected in the wastewater samples. For the viruses where epidemiological data on virus circulation in Belgium were available, these matched the wastewater data. The wastewater pretreatment that was used, optimized for viral enrichment, was as such not suited for the surveillance of bacteria and fungi.

**Conclusions:** Community circulation levels of respiratory viruses were well reflected in wastewater samples, indicating that wastewater-based epidemiology can be a valuable tool in the epidemiology and management of respiratory infections.

## INTRODUCTION

Wastewater-based epidemiology (WBE) can be used as a non-invasive tool to monitor the circulation of pathogens within a population. Since various body fluids are discharged into wastewater systems, the method includes but is not limited to detection of fecally excreted viruses and bacteria. Several countries use regular testing of wastewater in the surveillance of poliovirus [1]. Investigation of sewage water can also be applied to monitor the circulation of other enteric viruses such as norovirus, hepatitis A virus and hepatitis E virus [2], [3]. Since the start of the COVID-19 pandemic, wastewater has also been used to follow circulation levels of SARS-CoV-2 and it’s different variants of concern (VOC) [4], [5]. For this purpose, we have collected samples from a regional sewage treatment plant in the region of Leuven (Belgium) since December 2020. We found wastewater based surveillance to be an important objective indicator of SARS-CoV-2 circulation in the community, which can prove to be highly valuable in times of reducing testing capacity or lower willingness to test [6].

Acute respiratory tract infections (ARIs), including pneumonia, constitute a major disease burden worldwide, especially in young children and the elderly. According to the WHO, lower respiratory infections were the world’s most deadly communicable disease, ranked as the 4th leading cause of death globally in 2019 (so irrespective of the effect of the COVID-19 pandemic). Respiratory viruses, including influenza virus, EV/RV, RSV, HMPV, PIV, HBoV, Adv and HCoVs, are the leading cause of ARI’s in children as well as in older adults [7]–[10]. A large proportion of respiratory infections do not lead to hospitalization or medical care seeking, and therefore go undetected. To gain a better understanding of the burden of respiratory viruses on society, it is important to obtain detailed information on their circulation in the community. This knowledge could allow a better prediction and management of major outbreaks, and could guide physicians in their diagnosis. The current approach is usually based on reporting by a limited number of sentinel physicians and labs, which can lead to bias in the available data.

To allow quick diagnosis of respiratory pathogens in immunocompromised and/or critically ill patients with serious lower respiratory infection, a respiratory panel (RP) for simultaneous detection of 22 respiratory viruses (influenza A, influenza B, RSV, HMPV, PIV-1 to -4, Adv, HBoV, RV/EV, EV-D68, HPeV, HCoV-NL63, -229E, -OC43, -HKU-1, -SARS and – MERS, CMV, HSV-1 and -2) and 7 bacteria/fungi (*Mycoplasma pneumoniae, Coxiella burnetii, Chlamydia pneumoniae, Chlamydia psittaci, Streptococcus pneumoniae, Legionella pneumophila* and *Pneumocystis jiroveci*) was developed by the Department of Laboratory Medicine of the University Hospitals Leuven. Using this RP, we explored whether wastewater sampling can be used to monitor the circulation of respiratory pathogens at population level.

## METHODS

### Sampling

Wastewater samples were collected on average weekly, starting December 6^th^ 2020, from a large regional wastewater treatment plant (WWTP) in Leuven that treats municipal wastewater of approximately 115000 inhabitants. Samples (500 mL) of 24-hour composite influent wastewater were collected through a time-proportional automated sampler, which collects 50 mL of wastewater every 10 minutes in a large container. The samples were stored in a refrigerator at 4°C before transport to the laboratory.

### Viral TNA extraction

Virus concentration and filtration from the wastewater samples was performed as described previously [6]. Total nucleic acids (TNA) were extracted from 500 µL concentrated and filtered sewage samples or DNA/RNA free water (negative controls) using the MagMAX™ Viral/Pathogen Nucleic Acid Isolation Kit on Kingfisher Flex-96 (ThermoFisher Scientific, Europe). TNA were eluted in 50 µL elution buffer. Extractions were done in duplicate to obtain sufficient volume of elute for all the tests performed.

### Respiratory panel (RP)

An in-house developed RP consisting of 12 real-time multiplex PCRs was performed on a QuantStudio 7 (Thermo Fisher Scientific, Waltham, MA, USA) in 96 well plates. The end volume of each PCR reaction mix was 20 µL consisting of 5 µL of TNA, 5 µL of master mix (TaqMan Fast Virus Mix) and 10 µL of primer/probe mix. Primer and probe sequences and final concentrations are provided in Supplementary table 1. The temperature profile used was 50°C for 10 minutes; 95°C for 20 seconds; 45 cycles composed of 95°C for 3 seconds and 60°C for 30 seconds.

Specificity of this lab-developed RP was validated in a clinical context using External Quality Control (EQC) samples, virus cultures and clinical respiratory samples.

### Detection of PMMoV

Human fecal indicator pepper mild mottle virus (PMMoV) was analyzed as an internal extraction control, and to check for extensive differences in human waste input and/or rainwater infiltration, as described previously [6], [11].

## RESULTS AND DISCUSSION

Full RP results are listed in Supplementary table 2. The human fecal indicator PMMoV was detected in all wastewater samples with only limited variation in Ct values (Ct 22 to 26) (Fig. 1i), indicating there were no extensive differences in fecal input or wastewater dilution throughout the study. Although we did not determine the normalized pathogen concentrations using standard curves, the stable detection of PMMoV indicates that Ct values can be used as a proxy for semi-quantitative assessment of the pathogen load in our wastewater samples.

**Figure 1.**
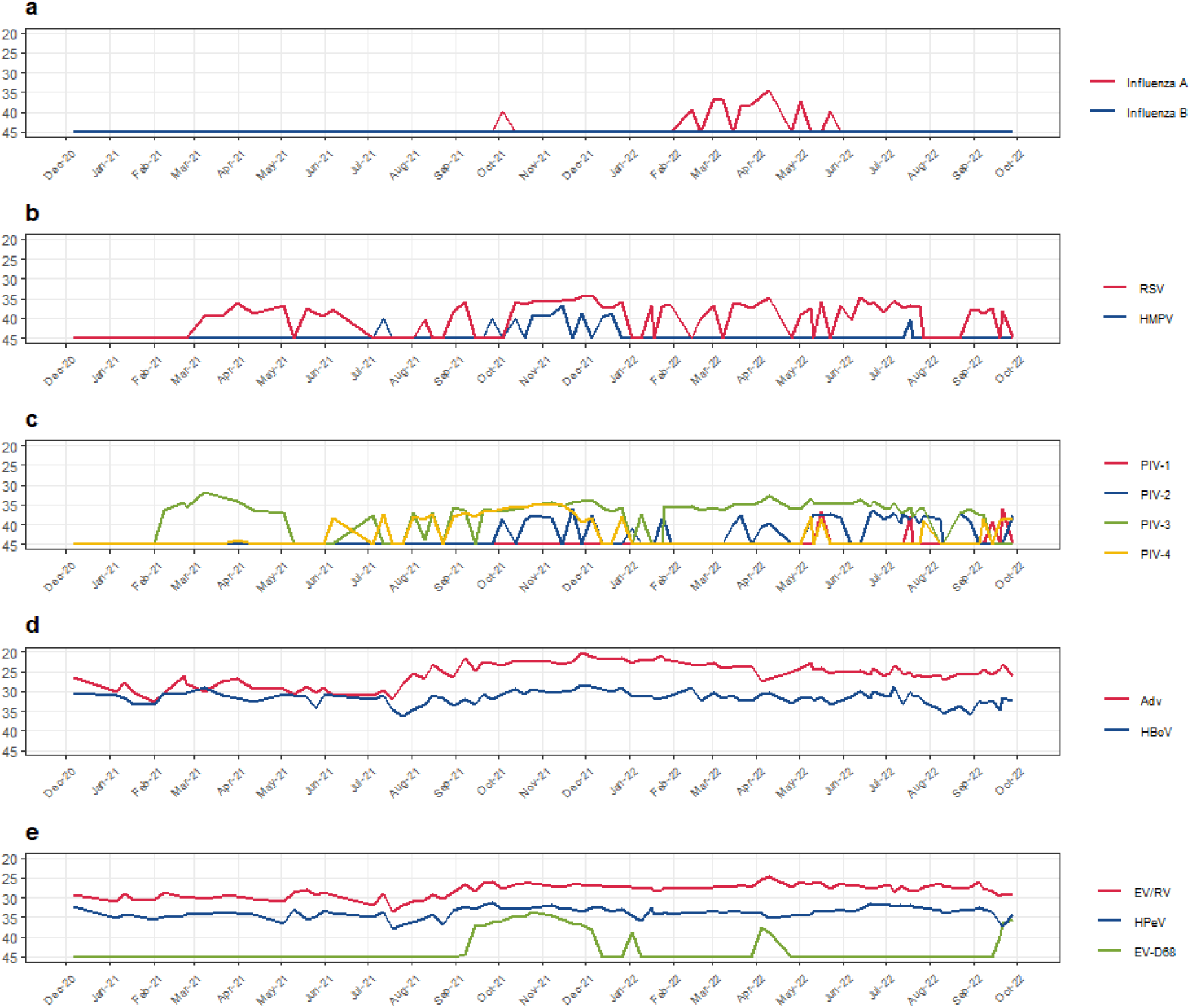
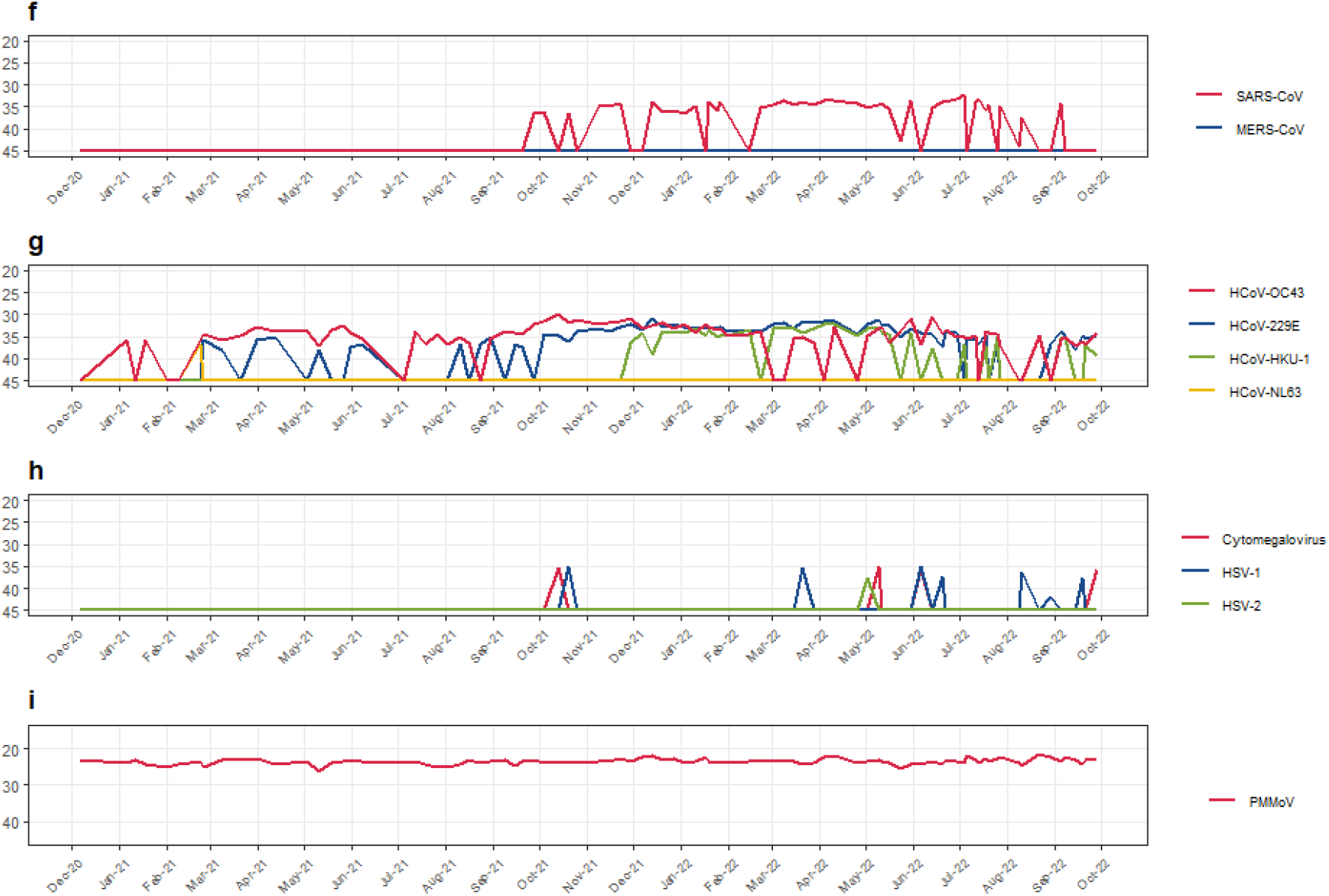
Respiratory viruses in wastewater (Aquafin, Leuven). Evolution of viruses detected by the respiratory panel from December 2020 to September 2022, as captured by the Ct value of targeted PCR assays performed on wastewater. Plots were generated using R and the ggplot2 package.

### Influenzavirus (Fig. 1a)

Whereas influenza B was not detected in any of the collected samples, influenza A was detected in only 1 sample in 2021 (dating from October 4^th^ 2021), and was repeatedly detected between mid Februari and mid May 2022. This is in line with the influenza data collected by the network of sentinel physicians and laboratories, coordinated by the national public health institute of Belgium Sciensano, who reported a very mild influenza season 2020-2021, and a late influenza season 2021-2022 between end of Februari 2022 and end of April 2022. Only influenza A was detected by the sentinel labs during the 2021-2022 season [12].

### RSV and HMPV (Fig. 1b)

RSV was detected in all but one wastewater sample collected between March 8^th^ and June 7^th^ 2021. Sporadic detection occurred during the summer of 2021. As of mid October 2021, RSV was continuously detected until end of December 2021, and discontinuously in January 2022. From the end of February 2022 until the end of July 2022, RSV was present in almost all wastewater samples. This is in accordance with the data from the sentinel labs reported by Sciensano, which indicated a late RSV season in 2019-2020 and a low, almost continuous RSV presence up to week 31 (beginning of August) during 2021-2022 [12]. Since the end of August 2022, RSV was again detectable in wastewater, but no infections have been reported by Sciensano yet in the weekly bulletin of week 40, which might be attributable to a delay in data reporting by the sentinel labs. HMPV was not detected in the wastewater samples during the winter season 2020-2021, and was found in one sample during the summer of 2021. From the end of September until the end of December 2021, HMPV was nearly continuously present in the wastewater samples. HMPV is not included in the respiratory virus report of Sciensano, so no data on the circulation of HMPV from sentinel labs in Belgium are available.

In the past, we have studied the prevalence and seasonality of respiratory viruses in respiratory samples of ARI patients in the University Hospitals of Leuven, from 2011-2016 [13]. These data showed that the HMPV season had a median onset around mid-December (week 50), peaked in the beginning of April (week 14.5) and lasted until May (week 21). For RSV, the median start of the epidemic season was in the late fall (week 44), with a peak on average at the beginning of December (week 48), and ending at the beginning of Februari (week 5). This does not match the circulation patterns we now see in the wastewater samples. As for other respiratory viruses, the circulation of RSV and HMPV was impacted by the nonpharmacological interventions such as extensive hand hygiene, mask-wearing, and social distancing that were implemented globally in response to the COVID-19 pandemic [14]–[16]. This can account for alterations in levels as well as timing of virus circulation.

### Parainfluenzaviruses (Fig. 1c)

Of the parainfluenzaviruses, PIV-1 was not detected in 2021 and only sporadically in 2022. PIV-2 was detected almost continuously during the fall of 2021, then more sporadically during the winter 2021-2022, and again almost continuously as off mid March 2022 until the end of August. PIV-3 was detected in all wastewater samples from the beginning of Februari 2021 until the beginning of May 2021. As of the beginning of August 2021, PIV-3 was again detectable in almost all wastewater samples up to the beginning of September 2022. PIV-4 was detectable in almost all samples between the August 2021 and December 2021, and again as of September 2022, and only sporadically in samples in between. In the data on respiratory pathogens detected by sentinel labs, shared by Sciensano, a peak in PIV circulation is visible between mid Februari 2021 and the beginning of April 2021. During the season 2021-2022, the PIV circulation was more spread out, with no clear peak being present [12]. Data on circulation of different PIV subtypes is not available from Sciensano. In our study on seasonality of respiratory viruses in ARI samples, we found that the 4 subtypes of PIV had their seasonal peaks at different moments during the year. Subtype 3 had the tendency to have its seasonal peaks around April/May, subtype 4 around December, while subtypes 1 and 2 did not present with a clear seasonal pattern [13]. According to our wastewater data, this seasonality was more or less maintained in 2021 (with a spring peak of PIV-3 and an (earlier) winter peak of PIV-4), but in 2022 PIV-3 was more continuously present.

### Adenovirus and bocavirus (Fig. 1d)

Both Adv and HBoV were detected consistently and in similar concentrations in all wastewater samples during the entire study. This is in accordance with the continuous Adv circulation in 2020-2021 and 2021-2022 reported by Sciensano, and with our previous study on ARI samples, were we found Adv infections to be present throughout the year, with a slight downfall during the Belgian summer season (week 25-38) [12], [13]. The circulation of HBoV is not reported by Sciensano, but its global prevalence has been reported from many countries, including Belgium [17]. To date, 4 genotypes of HBoV have been identified, all of which can be detected by the primers and probe located in the highly conserved NP-1 gene, used in our respiratory panel [18]. HBoV1 was mainly identified in respiratory samples from children with ARI [19], whereas HBoV2 is associated with clinical symptoms of patients with gastroenteritis [20]. All HBoV genotypes are known to be present in stool, and HBoV can frequently be detected in sewage samples [21]. AdV infections can also cause gastrointestinal symptoms, even when the primary site of involvement is the respiratory tract, and some serotypes (notably AdV-40 and -41) have an affinity for the gastrointestinal tract, with predominant symptoms of gastroenteritis or diarrhea [22]. The presence of HBoV and Adv in our wastewater samples is therefore likely linked to enteric infections rather than respiratory infections.

### Enterovirus/rhinovirus, EV-D68 and parechovirus (Fig. 1e)

Of the 15 classified species of enterovirus, EV A–D and RV A–C infect humans and are a frequent cause of respiratory infections. The fecal-oral route is the most common mode of EV transmission and the primary sites of replication are the gastrointestinal and respiratory tract [23]. Most infections are mild with fever and/or common cold symptoms, but some types, especially those belonging to EV A–D, may cause meningitis, encephalitis, paralysis, neonatal sepsis, myalgia, myocarditis, or exanthema. Of note, EV-D68 has recently been shown to cause large outbreaks of more severe disease with complications such as acute flaccid myelitis [24], [25]. Although EV/RV in general were detected continuously in the wastewater, EV-D68 detection was limited to early September 2021 until early December 2021, with the highest concentrations in October 2021. These data provide indications for an outbreak of EV-D68 in the region of Leuven during the fall of 2021, which is in line with the upsurge of EV-D68 infections in Europe as of September 2021, reported by the ECDC [26]. A comparison with the numbers of EV-D68 positive samples tested in the University Hospitals Leuven showed that a total of 20 patients were diagnosed with EV-D68 in 2021, all of which between October and December 2021 (unpublished data). There was a sporadic EV-D68 detection in January 2022, and a brief resurgence in April 2022. As of mid September 2022, EV-D68 was again detected in the wastewater samples, which could indicate the start of a new EV-D68 upsurge in the autumn of 2022.

Like EV/RV, HPeV was present in all samples during the entire study period. HPeV infections without specific symptoms are very common in children, but disease manifestations ranging from gastroenteritis and respiratory infections to neurological disease, particularly in neonates, have been described in association with HPeV [27]. Given their involvement in gastrointestinal infections, HPeVs are frequently detected in sewage samples, where their presence can be an early indicator of clinical cases, including CNS-infections [3], [28]. However, since HPeV were continuously detected in our wastewater samples, indicating a year-round circulation of HPeV in the community, this measurement does not seem useful as an early warning in our setting.

### Coronaviruses (Fig. 1f and 1g)

With the SARS-CoV assay in the respiratory panel, targeting a conserved region in the ORF1ab polyprotein gene to allow detection of both SARS-CoV-1 and -2, we did not detect SARS-CoV-2 until the end of September 2021 and only discontinuously thereafter. We also used the 2019-nCoV CDC EUA kit, targeting two regions of the N-gene, for detection of SARS-CoV-2 in all wastewater samples and were able to detect SARS-CoV-2 viral RNA at all measured timepoints, with viral RNA levels largely corresponding to the number of positive cases in Leuven [6]. This indicates that, although suitable in a clinical context, the universal RP SARS-CoV assay is not sensitive enough for environmental surveillance.

As expected, the highly pathogenic MERS-CoV was not detected at any timepoint, since MERS-CoV infections have not been reported in Europe for over 4 years now [29], [30].

Of the 4 endemic seasonal coronaviruses infecting humans, HCoV-NL-63 was only detected in one sample whereas HCoV-229E and HCoV-OC43 were present in 72% and 84% of all samples respectively. HCoV-HKU-1 was only detected as off the end of November 2021. These endemic coronaviruses are mainly associated with mild and self-limiting upper respiratory tract infections, accounting for about 15–30% of common cold cases, and normally circulate with annual peaks of infections in the winter months [31], [32]. Besides the occurrence in the respiratory tract, all endemic HCoVs can also be detected in stool samples but they do not seem to be a major cause of gastroenteritis, and this detection is most likely explained by the presence of ingested virus particles from the respiratory tract rather than resulting from productive replication in intestinal tissue [31]. In regions with a temperate climate, such as Belgium, the highest activity of endemic HCoVs is observed during the winter months [32]. As is the case for other respiratory viruses, their circulation pattern has been affected by contact restrictions and hygienic measures that were implemented in reaction to the COVID-19 outbreak, especially during the 1^st^ year after the emergence of SARS-CoV-2. This could account for the lower levels of endemic HCoV’s detected in the wastewater samples up to the spring of 2021.

### Herpesviruses (Fig. 1h)

HSV can cause influenza like illness, but also meningitis and encephalitis. CMV is a ubiquitous pathogen mainly infecting children, with childhood infections usually going unnoticed whereas infections in adults can be more severe, especially in the immunocompromised. There are only limited data available on the presence of human herpesviruses in wastewater, especially for HSV [33]. In our study, HSV-1 and CMV were only sporadically detected, each at 4 scattered timepoints, and HSV-2 was only detected in a single wastewater sample.

### Bacteria and fungi

In view of SARS-CoV-2 and polio surveillance, the current method was developed for optimal retrieval of viral material, by including filtration using a 1 µm syringe filter. The drawback of this viral enrichment is that the majority of bacterial and fungal material is lost prior to extraction. Although we were able to detect bacteria/fungi in some of the samples, we consider this method unsuited to confidently detect their presence in wastewater.

## CONCLUSION

By using the RP on our sample collection spanning almost 2 years, we were able to detect the presence and seasonal variation for most of the tested respiratory viruses. This indicates that wastewater sampling could provide a means to monitor the circulation of respiratory pathogens at population level, and provide an early warning signal in case of local upsurges. By using wastewater samples, the inherent bias posed by data reporting by limited numbers of sentinel labs can be circumvented.

## Data Availability

All data produced in the present study are available upon reasonable request to the authors

https://www.sciensano.be/en

---

**Supplementary table 1:**
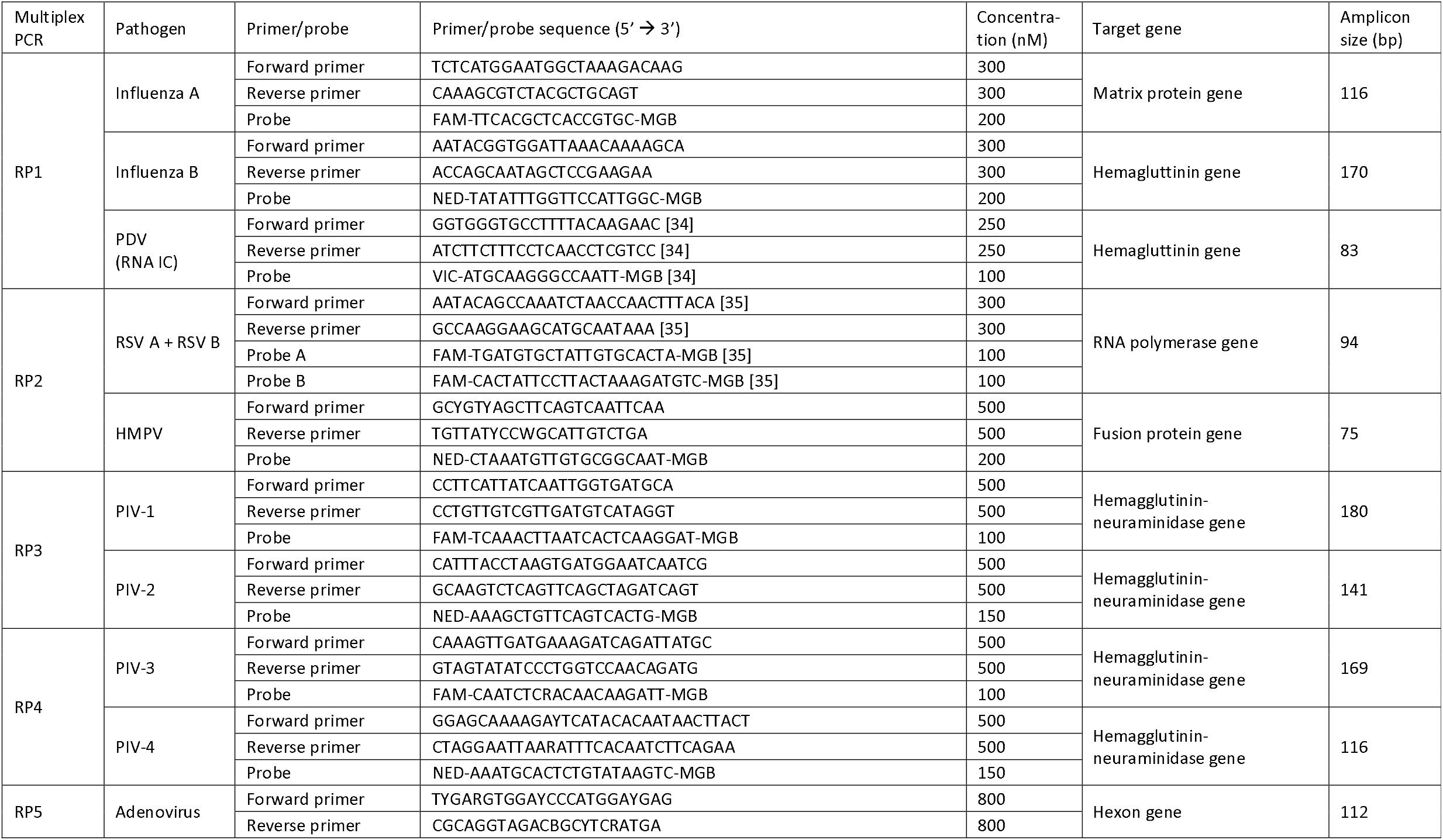
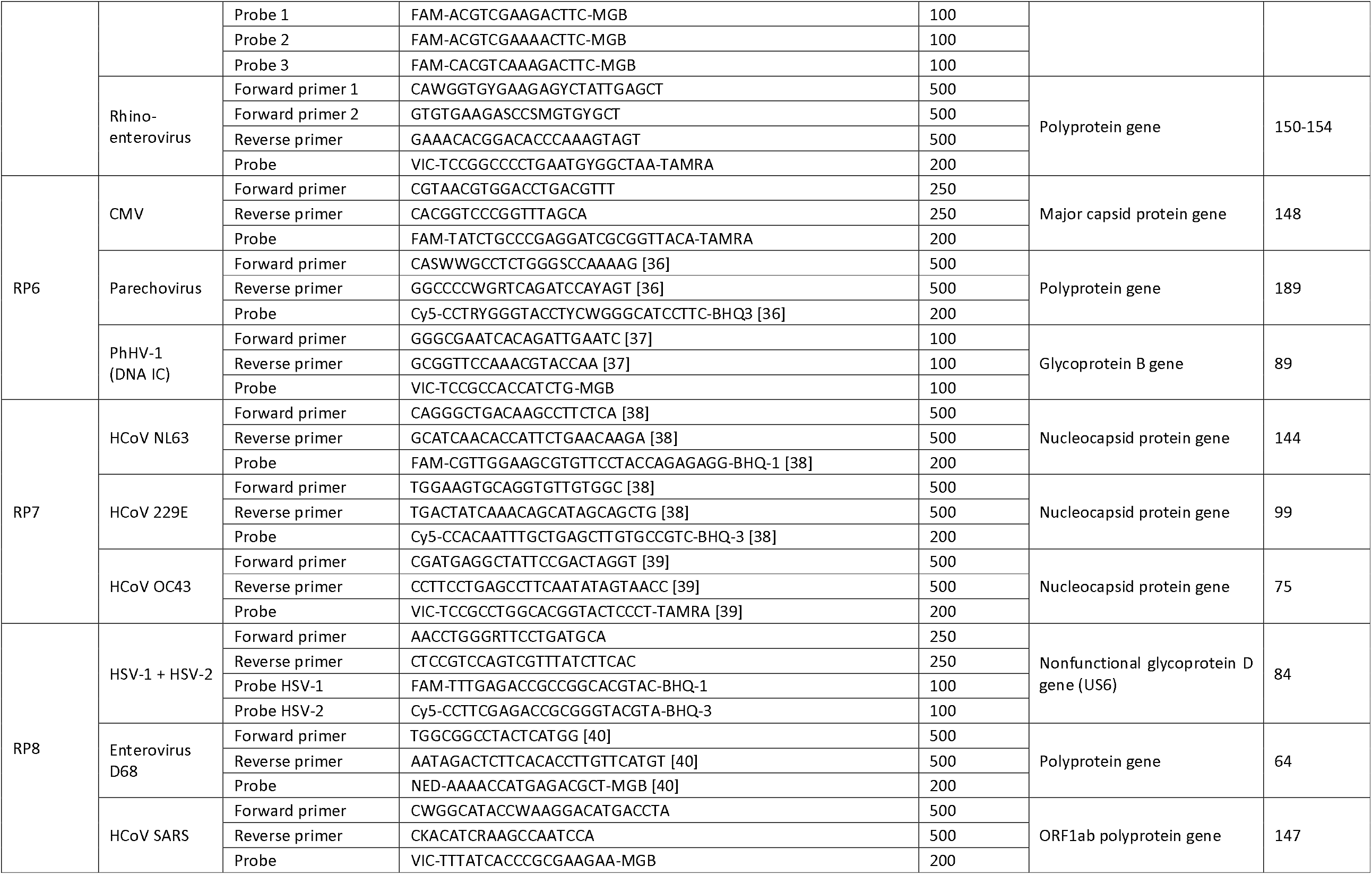
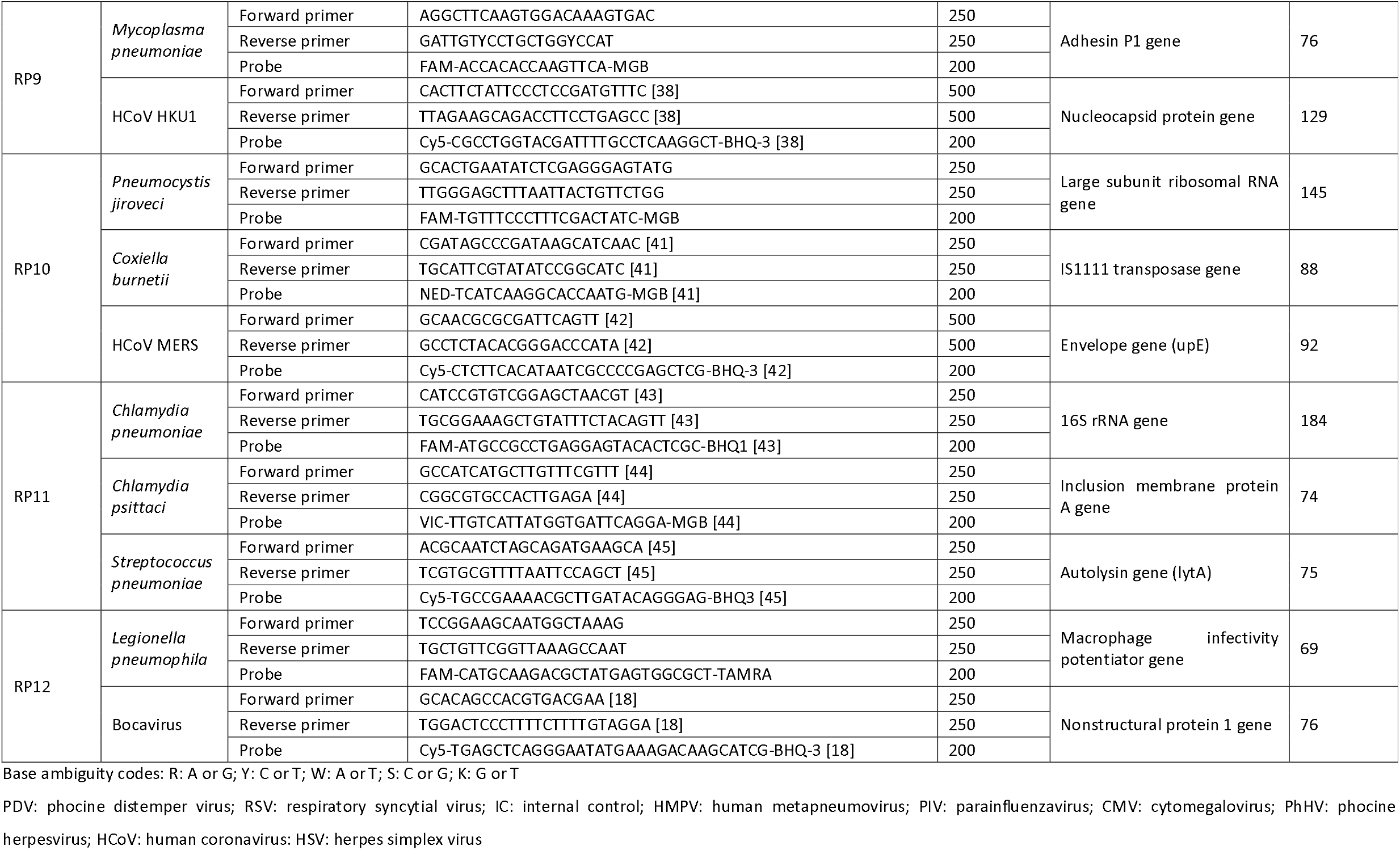
Primers and probes used in the 12 multiplex PCR reactions of the respiratory panel.

**Supplementary table 2:** Ct values of pathogens detected by the respiratory panel in wastewater samples (Aquafin, Leuven) at different timepoints.

|  | Influenza A | Influenza B | RSV | HMPV | PIV-1 | PIV-2 | PIV-3 | PIV-4 | Adv | HBov | EV/RV | EV-D68 | HPeV | SARS-CoV | MERS-CoV | HCov-NL63 | HCov-229E | HCov-OC43 | HCov-HKU-1 | HSV-1 | HSV-2 | Cytomegalovirus | PMMoV |
| --- | --- | --- | --- | --- | --- | --- | --- | --- | --- | --- | --- | --- | --- | --- | --- | --- | --- | --- | --- | --- | --- | --- | --- |
| 6/12/20 |  |  |  |  |  |  |  |  | 27 | 30 | 29 |  | 32 |  |  |  |  |  |  |  |  |  | 23 |
| 5/01/21 |  |  |  |  |  |  |  |  | 30 | 31 | 31 |  | 35 |  |  |  |  | 36 |  |  |  |  | 24 |
| 11/01/21 |  |  |  |  |  |  |  |  | 28 | 32 | 29 |  | 34 |  |  |  |  |  |  |  |  |  | 23 |
| 18/01/21 |  |  |  |  |  |  |  |  | 30 | 33 | 31 |  | 35 |  |  |  |  | 36 |  |  |  |  | 24 |
| 1/02/21 |  |  |  |  |  |  |  |  | 33 | 33 | 31 |  | 36 |  |  |  |  |  |  |  |  |  | 25 |
| 8/02/21 |  |  |  |  |  |  | 36 |  | 30 | 31 | 29 |  | 35 |  |  |  |  |  |  |  |  |  | 24 |
| 22/02/21 |  |  |  |  |  |  | 34 |  | 26 | 31 | 30 |  | 34 |  |  | 37 |  | 36 |  |  |  |  | 23 |
| 24/02/21 |  |  |  |  |  |  | 36 |  | 28 | 30 | 30 |  | 34 |  |  |  | 36 | 35 |  |  |  |  | 25 |
| 8/03/21 |  |  | 39 |  |  |  | 32 |  | 30 | 29 | 30 |  | 34 |  |  |  | 38 | 36 |  |  |  |  | 23 |
| 21/03/21 |  |  | 39 |  |  |  | 33 |  | 27 | 31 | 30 |  | 33 |  |  |  |  | 35 |  |  |  |  | 23 |
| 31/03/21 |  |  | 36 |  |  |  | 34 | 44 | 27 | 32 | 30 |  | 34 |  |  |  | 36 | 33 |  |  |  |  | 23 |
| 12/04/21 |  |  | 39 |  |  |  | 37 |  | 30 | 33 | 30 |  | 34 |  |  |  | 35 | 34 |  |  |  |  | 25 |
| 3/05/21 |  |  | 37 |  |  |  | 37 |  | 29 | 31 | 31 |  | 36 |  |  |  |  | 34 |  |  |  |  | 24 |
| 10/05/21 |  |  |  |  |  |  |  |  | 31 | 31 | 29 |  | 33 |  |  |  | 38 | 37 |  |  |  |  | 26 |
| 19/05/21 |  |  | 37 |  |  |  |  |  | 29 | 32 | 28 |  | 35 |  |  |  |  | 34 |  |  |  |  | 24 |
| 26/05/21 |  |  | 39 |  |  |  |  |  | 30 | 34 | 29 |  | 35 |  |  |  |  | 32 |  |  |  |  | 24 |
| 31/05/21 |  |  | 39 |  |  |  |  |  | 29 | 31 | 29 |  | 33 |  |  |  | 38 | 34 |  |  |  |  | 23 |
| 7/06/21 |  |  | 38 |  |  |  |  | 38 | 31 | 31 | 29 |  | 34 |  |  |  | 37 | 35 |  |  |  |  | 24 |
| 5/07/21 |  |  |  |  |  |  | 38 |  | 31 | 32 | 32 |  | 35 |  |  |  |  |  |  |  |  |  | 24 |
| 12/07/21 |  |  |  | 40 |  |  |  | 37 | 30 | 31 | 29 |  | 34 |  |  |  |  | 34 |  |  |  |  | 23 |
| 19/07/21 |  |  |  |  |  |  |  |  | 32 | 35 | 34 |  | 38 |  |  |  |  | 37 |  |  |  |  | 24 |
| 26/07/21 |  |  |  |  |  |  |  |  | 28 | 36 | 32 |  | 37 |  |  |  |  | 35 |  |  |  |  | 25 |
| 2/08/21 |  |  |  |  |  |  | 37 | 38 | 25 | 35 | 31 |  | 36 |  |  |  |  | 37 |  |  |  |  | 25 |
| 10/08/21 |  |  | 40 |  |  |  | 44 | 39 | 27 | 33 | 31 |  | 35 |  |  |  | 37 | 35 |  |  |  |  | 24 |
| 16/08/21 |  |  |  |  |  |  | 37 | 37 | 23 | 31 | 29 |  | 34 |  |  |  |  | 36 |  |  |  |  | 23 |
| 23/08/21 |  |  |  |  |  |  |  |  | 25 | 32 | 30 |  | 37 |  |  |  | 37 |  |  |  |  |  | 24 |
| 30/08/21 |  |  | 39 |  |  |  | 36 | 38 | 26 | 34 | 29 |  | 33 |  |  |  | 35 | 35 |  |  |  |  | 23 |
| 8/09/21 |  |  | 36 |  |  |  | 36 | 37 | 22 | 32 | 27 | 44 | 32 |  |  |  |  | 34 |  |  |  |  | 23 |
| 15/09/21 |  |  |  |  |  |  |  | 38 | 25 | 33 | 28 | 37 | 33 |  |  |  | 37 | 34 |  |  |  |  | 25 |
| 20/09/21 |  |  |  |  |  |  | 37 | 37 | 22 | 31 | 26 | 37 | 32 |  |  |  | 37 | 34 |  |  |  |  | 23 |
| 27/09/21 |  |  |  | 40 |  |  | 36 | 37 | 23 | 32 | 26 | 36 | 31 | 36 |  |  |  | 32 |  |  |  |  | 23 |
| 4/10/21 | 40 |  |  |  |  | 39 | 37 | 36 | 23 | 31 | 28 | 36 | 33 | 36 |  |  | 34 | 32 |  |  |  |  | 24 |
| 13/10/21 |  |  | 36 | 40 |  |  | 36 | 35 | 22 | 29 | 27 | 35 | 33 |  |  |  | 35 | 30 |  |  |  | 35 | 23 |
| 20/10/21 |  |  | 36 |  |  | 39 | 36 | 36 | 23 | 31 | 26 | 35 | 33 | 37 |  |  | 36 | 32 |  | 35 |  |  | 24 |
| 25/10/21 |  |  | 36 | 39 |  | 38 | 35 | 35 | 22 | 29 | 26 | 34 | 32 |  |  |  | 34 | 31 |  |  |  |  | 24 |
| 8/11/21 |  |  | 36 | 39 |  | 38 | 34 | 35 | 22 | 30 | 27 | 35 | 32 | 35 |  |  | 33 | 32 |  |  |  |  | 23 |
| 15/11/21 |  |  | 36 | 37 |  |  | 36 | 35 | 23 | 30 | 27 | 36 | 33 | 35 |  |  | 34 | 32 |  |  |  |  | 24 |
| 22/11/21 |  |  | 35 |  |  | 36 | 34 | 37 | 23 | 30 | 27 | 37 | 33 | 34 |  |  | 33 | 31 |  |  |  |  | 23 |
| 29/11/21 |  |  | 34 | 39 |  |  | 34 | 39 | 20 | 28 | 27 | 37 | 33 |  |  |  | 32 | 31 | 36 |  |  |  | 24 |
| 6/12/21 |  |  | 34 |  |  | 38 | 34 | 38 | 21 | 29 | 27 | 38 | 33 |  |  |  | 34 | 33 | 34 |  |  |  | 22 |
| 13/12/21 |  |  | 37 | 40 |  |  | 36 |  | 22 | 30 | 27 |  | 33 | 34 |  |  | 31 | 33 | 39 |  |  |  | 22 |
| 20/12/21 |  |  | 38 | 39 |  |  | 37 |  | 22 | 30 | 27 |  | 34 | 36 |  |  | 33 | 32 | 34 |  |  |  | 23 |
| 27/12/21 |  |  | 36 |  |  |  | 36 | 38 | 21 | 29 | 27 |  | 33 | 36 |  |  | 32 | 33 | 34 |  |  |  | 23 |
| 3/01/22 |  |  |  |  |  | 41 |  |  | 23 | 31 | F | 39 | 35 | 36 |  |  | 33 | 32 | 34 |  |  |  | 24 |
| 10/01/22 |  |  |  |  |  |  | 37 |  | 22 | 31 | 28 |  | 36 | 35 |  |  | 33 | 34 | 33 |  |  |  | 24 |
| 17/01/22 |  |  | 37 |  |  |  |  |  | 22 | 31 | 28 |  | 33 |  |  |  | 33 | 32 | 35 |  |  |  | 23 |
| 19/01/22 |  |  |  |  |  |  |  |  | 22 | 32 | 28 |  | 34 | 34 |  |  | 33 | 33 | 34 |  |  |  | 24 |
| 24/01/22 |  |  | 37 |  |  | 39 |  |  | 21 | 32 | 28 |  | 34 | 36 |  |  | 33 | 33 | 35 |  |  |  | 24 |
| 31/01/22 |  |  | 37 |  |  |  | 36 |  | 22 | 31 | 28 |  | 34 | 36 |  |  | 34 | 35 | 35 |  |  |  | 24 |
| 26/01/22 |  |  | 37 |  |  | 39 | 35 |  | 22 | 32 | 27 |  | 33 | 34 |  |  | 33 | 33 | 35 |  |  |  | 24 |
| 14/02/22 | 39 |  |  |  |  |  | 35 |  | 23 | 29 | 28 |  | 34 |  |  |  | 34 | 34 | 33 |  |  |  | 24 |
| 21/02/22 |  |  | 40 |  |  |  | 36 |  | 24 | 32 | 28 |  | 34 | 35 |  |  | 34 | 34 |  |  |  |  | 23 |
| 2/03/22 | 37 |  | 37 |  |  |  | 36 |  | 23 | 31 | 27 |  | 33 | 34 |  |  | 32 |  | 33 |  |  |  | 23 |
| 8/03/22 | 37 |  |  |  |  |  | 36 |  | 24 | 32 | 27 |  | 34 | 34 |  |  | 32 |  | 33 |  |  |  | 24 |
| 15/03/22 |  |  | 36 |  |  | 40 | 35 |  | 24 | 31 | 28 |  | 33 | 35 |  |  | 33 | 35 | 33 |  |  |  | 24 |
| 21/03/22 | 38 |  | 36 |  |  | 38 | 35 |  | 24 | 32 | 27 |  | 33 | 34 |  |  | 32 | 35 | 34 | 36 |  |  | 24 |
| 28/03/22 | 38 |  | 37 |  |  |  | 35 |  | 23 | 32 | 27 |  | 34 | 34 |  |  | 32 | 36 | 33 |  |  |  | 24 |
| 4/04/22 | 36 |  | 36 |  |  | 41 | 34 |  | 27 | 31 | 25 | 37 | 34 | 33 |  |  | 31 |  | 32 |  |  |  | 22 |
| 10/04/22 | 35 |  | 35 |  |  | 40 | 33 |  | 27 | 30 | 25 | 39 | 35 | 34 |  |  | 32 | 33 | 32 |  |  |  | 22 |
| 25/04/22 |  |  |  |  |  |  | 36 |  | 25 | 33 | 28 |  | 34 | 34 |  |  | 34 |  | 35 |  |  |  | 24 |
| 2/05/22 | 37 |  | 39 |  |  |  | 36 |  | 24 | 32 | 26 |  | 35 | 35 |  |  | 32 | 35 | 33 |  | 38 |  | 23 |
| 9/05/22 |  |  | 38 |  |  | 39 | 33 | 38 | 23 | 31 | 27 |  | 34 | 35 |  |  | 31 | 33 | 33 |  |  | 35 | 23 |
| 11/05/22 |  |  |  |  |  | 37 | 35 |  | 24 | 32 | 26 |  | 34 | 34 |  |  | 33 | 33 | 34 |  |  |  | 24 |
| 16/05/22 |  |  | 36 |  | 37 | 38 | 35 | 39 | 24 | 31 | 26 |  | 34 | 36 |  |  | 32 | 36 | 35 |  |  |  | 24 |
| 23/05/22 | 40 |  |  |  |  | 37 | 35 |  | 26 | 33 | 28 |  | 33 | 43 |  |  | 35 | 34 |  |  |  |  | 25 |
| 30/05/22 |  |  | 37 |  |  | 38 | 35 |  | 25 | 32 | 26 |  | 33 | 34 |  |  | 33 | 31 | 34 |  |  |  | 24 |
| 6/06/22 |  |  | 40 |  |  |  | 35 |  | 25 | 31 | 27 |  | 34 |  |  |  | 34 | 37 |  | 35 |  | 36 | 24 |
| 13/06/22 |  |  | 35 |  |  |  | 34 |  | 25 | 30 | 27 |  | 33 | 35 |  |  | 34 | 31 | 38 |  |  |  | 24 |
| 20/06/22 |  |  | 36 |  |  | 37 | 35 |  | 26 | 32 | 28 |  | 32 | 34 |  |  | 37 | 35 |  | 37 |  |  | 24 |
| 22/06/22 |  |  | 36 |  |  | 36 | 34 |  | 24 | 30 | 28 |  | 32 | 34 |  |  | 34 | 34 |  |  |  |  | 23 |
| 29/06/22 |  |  | 39 |  |  | 39 | 36 |  | 26 | 31 | 27 |  | 31 | 33 |  |  | 34 | 35 |  |  |  |  | 24 |
| 4/07/22 |  |  | 37 |  |  | 38 | 36 |  | 25 | 31 | 27 |  | 32 | 32 |  |  |  | 35 | 37 |  |  |  | 24 |
| 6/07/22 |  |  | 35 |  |  | 37 | 34 |  | 23 | 29 | 29 |  | 32 |  |  |  | 36 | 35 |  |  |  |  | 22 |
| 11/07/22 |  |  | 37 |  |  | 40 | 37 |  | 25 | 32 | 27 |  | 32 | 34 |  |  | 36 | 35 |  |  |  |  | 23 |
| 13/07/22 |  |  | 37 |  |  | 37 | 37 |  | 26 | 33 | 28 |  | 32 | 33 |  |  | 37 |  |  |  |  |  | 24 |
| 18/07/22 |  |  | 37 | 40 | 38 | 39 | 36 |  | 25 | 30 | 28 |  | 32 | 36 |  |  | 34 | 34 | 37 |  |  |  | 23 |
| 20/07/22 |  |  | 37 |  |  | 38 | 38 |  | 26 | 32 | 28 |  | 32 | 34 |  |  |  | 34 |  |  |  |  | 24 |
| 25/07/22 |  |  | 37 |  |  | 40 | 36 |  | 26 | 31 | 28 |  | 33 |  |  |  | 37 | 34 | 35 |  |  |  | 23 |
| 27/07/22 |  |  |  |  |  | 37 | 36 | 39 | 26 | 33 | 27 |  | 33 | 35 |  |  |  | 37 |  |  |  |  | 22 |
| 8/08/22 |  |  |  |  |  | 39 |  |  | 26 | 35 | 26 |  | 33 | 44 |  |  |  |  |  |  |  |  | 24 |
| 10/08/22 |  |  |  |  |  |  |  |  | 27 | 36 | 27 |  | 34 | 38 |  |  |  |  |  | 36 |  |  | 25 |
| 22/08/22 |  |  |  |  |  | 37 | 37 |  | 25 | 34 | 28 |  | 33 |  |  |  |  | 35 |  |  |  |  | 22 |
| 29/08/22 |  |  | 38 |  |  | 39 | 36 |  | 26 | 36 | 28 |  | 33 |  |  |  | 37 |  |  | 42 |  |  | 22 |
| 5/09/22 |  |  | 38 |  |  |  | 38 |  | 25 | 32 | 26 |  | 33 | 34 |  |  | 34 | 36 | 36 |  |  |  | 24 |
| 7/09/22 |  |  | 39 |  |  |  | 37 | 39 | 25 | 33 | 28 |  | 33 |  |  |  | 35 | 35 | 35 |  |  |  | 22 |
| 14/09/22 |  |  | 38 |  | 39 |  |  |  | 26 | 32 | 29 |  | 34 |  |  |  | 38 | 37 |  |  |  |  | 23 |
| 19/09/22 |  |  |  |  |  |  |  | 39 | 25 | 34 | 30 | 40 | 36 |  |  |  | 35 | 36 |  | 38 |  |  | 24 |
| 21/09/22 |  |  | 38 |  | 36 |  |  | 39 | 23 | 32 | 29 | 37 | 37 |  |  |  | 35 | 37 | 37 |  |  |  | 23 |
| 28/09/22 |  |  |  |  |  | 38 |  | 38 | 26 | 32 | 29 | 36 | 34 |  |  |  | 35 | 34 | 39 |  |  | 36 | 23 |

